# Can Electronic care planning using AI Summarization Yield equal Documentation Quality? (EASY eDocQ)

**DOI:** 10.1101/2025.01.22.25320982

**Authors:** David A. Dorr, Nicole G. Weiskopf, Jean M Sabile, Emma Young, Dylan Mathis, Samantha Weller, Jacob Alex, Kimberly Doerr, Steven Bedrick

**Affiliations:** Oregon Health & Science University, Portland, OR

## Abstract

**Importance:** Data, information and knowledge in health care has expanded exponentially over the last 50 years, leading to significant challenges with information overload and complex, fragmented care plans. Generative AI has the potential to facilitate summarization and integration of knowledge and wisdom to through rapid integration of data and information to enable efficient care planning.

**Objective:** Our objective was to understand the value of AI generated summarization through short synopses at the care transition from hospital to first outpatient visit.

**Design:** Using a de-identified data set of recently hospitalized patients with multiple chronic illnesses, we used the data-information-knowledge-wisdom framework to train clinicians and an open-source generative AI Large Language Model system to produce summarized patient assessments after hospitalizations. Both sets of synopses were judged blinded in random order by clinician judges.

**Participants:** De-identified patients had multiple chronic conditions and a recent hospitalization. Raters were physicians at various levels of training.

**Main outcome:** Accuracy, succinctness, synthesis and usefulness of synopses using a standardized scale with scores > 80% indicating success.

**Results:** AI and clinicians summarized 80 patients with 10% overlap. In blinded trials, AI synopses were rated as useful 75% of the time versus 76% for human generated ones. AI had lower succinctness ratings for the Data synopsis task (55-67%) versus human (84-86%). For accuracy and synthesis, AI had near equal or better scores in other domains (AI: 72%-79%, humans: 68%-84%), with best scores from AI in Wisdom. Interrater agreement was moderate, indicating different preferences for synopsis content, and did not vary between AI and human-created synopses.

**Discussion:** AI-created synopses that were nearly equivalent to human-created ones; they were slightly longer and did not always synthesize individual data elements compared to humans. Given their rapid introduction into clinical care, our framework and protocol for evaluation of these tools provides strong benchmarking capabilities for developers and implementers.

**Key Points:** *Question:* Can a Generative AI Large Language Model be trained to generate accurate and useful patient synopses through chart summarization for use in outpatient settings after hospital discharge?

*Findings:* Using a Data-Information-Knowledge-Wisdom framework, clinicians and an open-source AI system were trained to summarize charts; these synopses were rated blindly using a standardized index. Synopses from the AI were rated as useful 75% of the time versus 76% for human generated ones, AI synopses scored highest in Wisdom for accuracy and synthesis. Interrater agreement was moderate but did not vary between AI and human.

*Meaning:* This study provides a concrete, replicable protocol for benchmarking LLM summarization outputs and demonstrates general equivalence to human-created synopses for outpatient use after care transitions.

## Introduction

Health care delivery has become a data and knowledge intensive exercise, especially when caring for those with complex conditions. For instance, persons living with Multiple Chronic Conditions utilize more health care and suffer exponentially worse outcomes than those with one or zero conditions. This is especially true of those with five or more conditions, who have ninety times the risk of hospitalization, are on an average of eight long term prescriptions, and see twelve different providers per year.^1-5^ Managing the needs of these patients in the context of outpatient visits is nearly impossible – estimates are that addressing the acute, preventive, and chronic illness needs of the patients for a single primary care physician would take 26 hours a day.^6^ Exacerbating this problem in the United States are the requirements for medical documentation. Billing expectations have led to notes that are highly redundant and up four-times longer than elsewhere in the world.^7-9^ These expectations also necessitate more frequent documentation by care team members, where information must be duplicated across different notes to facilitate billing, leading to substantial bloat and making concise communication difficult.

A major focus of ambulatory care teams and our previous research is care planning^10,11^, which can be defined as the proactive co-creation of an integrated treatment plan that incorporates patient health needs, values, and preferences. Care planning requires significant time to forage through data, derive current and relevant information, incorporate knowledge of guideline-based recommendations, and leverage the wisdom and experience of the care team and patient into the process. The care planning process is often summarized in the assessment and plan in notes, where a brief overall summary of the key information and recommendations are included. Care planning is challenging for several reasons, including the burdens of finding and extracting relevant information from clinical data, summarizing that information in a meaningful way, and incorporating appropriate clinical recommendations based on patient needs and preferences. Time to create care plans is substantial, and errors from missed information are common. These burdens have led to substantial burn-out amongst ambulatory care teams, with 30% of clinicians reporting burn-out and 50% of these indicating the EHR worsened their burn-out.^12^

Novel solutions exist. Generative AI models built on large textual corpora (called Large Language Models, or LLMs) generally use large neural networks (sometimes with more than 10^26^ elements) to predict what text should come next. Studies have shown benefits in auto-composing responses to patient questions, generating differential diagnoses from cases, and generating notes from ambient recordings.^13^ As a baseline, we sought to understand the ability of a set of advanced but generalized LLMs to produce clinically meaningful summaries from patient data with complex needs.

## Background

As described above, the major challenges involved in care planning can be broken into three broad steps: information identification and extraction, summarization, and appropriate recommendations. For care planning, one framework for summarization can be defined through the data, information, knowledge, and wisdom model.^14-17^ In this model, data are individual elements relevant to care planning; information is data with patient context, such as a condition (diabetes) with A1c and diet and activity self-management; knowledge incorporates evidence about diagnoses, prognosis, monitoring and effective treatments; and wisdom incorporates values, preferences, and needs that allow a care plan to be most aligned with the wishes of the patient.

### Identification, extraction, and prioritization of clinical information

EHRs have received substantial criticism for their cognitive burden and lack of usability in data finding and navigating, requiring significant expertise to gain full efficiency without substantial cognitive load. Data are fragmented, both within electronic systems and between settings of care. Gathering these data together – referred to as ‘foraging’ in the vernacular – takes substantial time and effort. Often a step known as ‘scrubbing’ helps to identify key information before outpatient visits to streamline care; observational studies of providers before, during, and after visits demonstrate a substantial (up to an average of 12 minutes per patient) ^18,19^ amount of time gathering information to understand what to ask; how to act on what has been learned; and assure themselves that these actions were complete and appropriate. Prioritization of these data is crucial: the volume of information available far outstrips the ability of an individual to consume and process it^20^, leading to substantial errors.^21^

### Summarization of information to provide patients with knowledge

A common aspect of training in medicine is to summarize key findings from complex patients in brief, clearly understood statements. Chart summarizations, synopses, and other documents serve to make it easier to quickly understand trends, new concerns and data, and other relevant information. ^22^ LLMs now achieve very high performance with providing clinical summary and a progressively lower barrier for utilization.^23-25^ However, the ability to implement flexible and adaptive summarizations is still challenging, requiring extensive technical experience.

Pivovarov and Elhadad summarized the work up to 2015, identifying significant work in removing redundancy, determining salience, and taking advantage of clinical knowledge; they highlight slow uptake of these systems.^26^ In 2017, Pivovarov published an extraction tool that identified salient concepts and presented them to quality metric summarizers, highlighting a 50% reduction in summarization time.^27^ In 2019, Liang, Tsou, and Poddar created a machine learning disease specific text summarization, achieving moderate agreement with human annotators (AUC .67-.74).^28^ Recent, cutting-edge word representation models, such as BERT, have been shown to be effective at biomedical text mining via extensive training on biomedical corpora, now termed bioBERT.^29-31^ More recently, GPT-4 was utilized to compare LLM clinical summaries to medical expert review and demonstrated up to 36% of AI-generated summaries were superior to medical experts and up to 45% of AI summaries were deemed equivalent.^23^ As summarization technology improves, benchmarking has also been developed to establish a standard on the efficacy of NLM models in biomedical data summarization.^32^

While promising, prior work has failed to address the core outpatient summarization steps that are required for care planning. With the advent of Generative AI tools, it may be possible to better replicate these steps. We performed a structured evaluation of these steps in order to both set a benchmark for LLM performance as well as share a structured evaluation protocol.

## Methods

### Overview

We conducted a non-inferiority trial to determine if a generalist LLM (Llama-8-3b)^33^ was able to perform comparably to human clinicians in the generation of actionable clinical synopses of discharge notes and associated structured data. Specifically, these synopses were intended to guide outpatient care for patients with Multiple Chronic Conditions after discharge from the hospital. We used the Data-Information-Knowledge-Wisdom framework to model different approaches to synopsis generation. The quality of clinician- and LLM-generated synopses were judged and then compared using a blinded design informed by the Physician Documentation Quality Instrument (PDQI).^34^ The study design is summarized in Figure 1. The OHSU Institutional Review Board determined this work to be not human subjects research (STUDY00026857).

**Figure 1.**
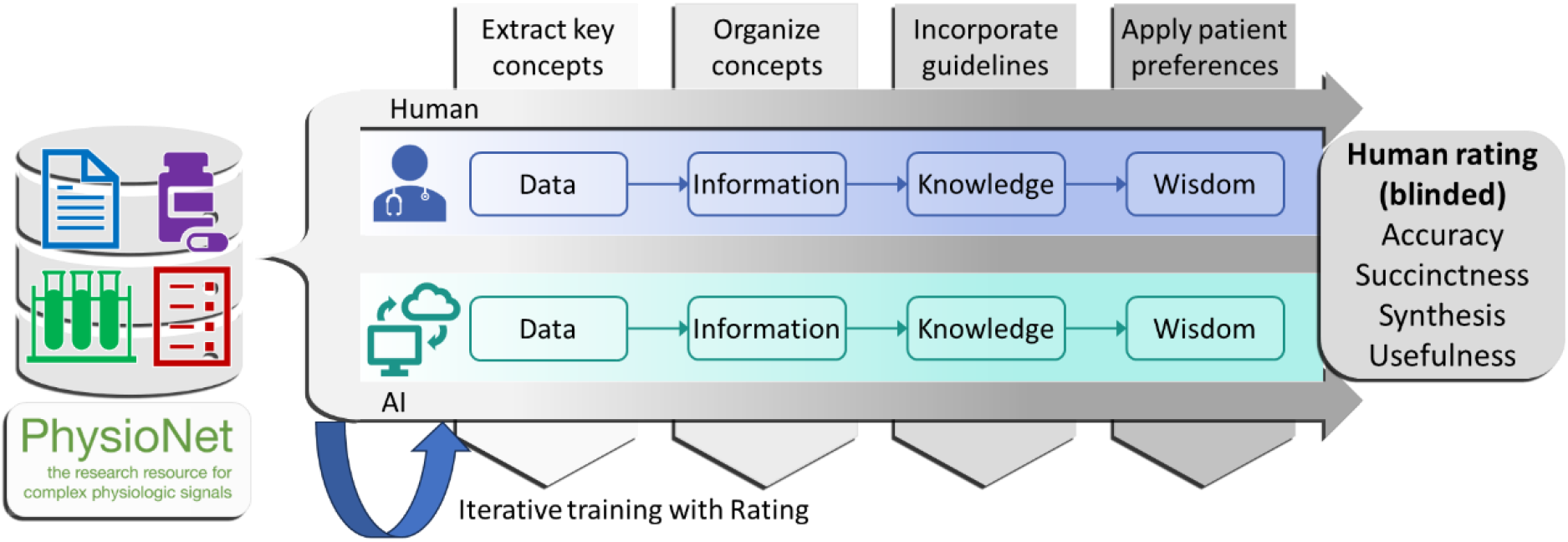
Study framework and design

### Data

After completing all relevant agreements and trainings on PhysioNet^35^ (see appendix), we used de-identified patients from the MIMIC-III database with multiple chronic conditions and significant outpatient follow-up needs. Based on our work in the eCarePlan project, we selected patients with a recent hospitalization with multiple chronic conditions for whom care planning was challenging due to the complexity of their needs. For multiple chronic conditions, we used a subset of conditions from the Chronic Condition Warehouse (see Appendix Table 1).^36^ We also leveraged the CLIP study^37^, which abstracted action items from discharge summaries for outpatient follow-up; patients who were part of the study and were discharged to home or home health were included.

### Synopsis Authoring and Prompt Engineering

As noted above, the generalist LLM and our human authors were asked to generate four synopses of each assigned discharge note and associated structured data, following the D-I-K-W framework. The high-level steps were to: identify and summarize key observations, conditions, treatments, and needs (data); contextualize these data (information); integrate evidence-based guidelines (knowledge); and incorporate social and behavioral needs with overall health for person-centered care planning (wisdom). The protocol and instructions for the synopsis authoring task provided to clinicians and final prompts for the LLM are available as supplemental materials. The LLM was not provided with additional documents for knowledge, instead relying on its generalist training.

An iterative process was used to develop LLM prompts and instructions for human authors. First, high-quality synopses adhering to the DIKW framework were written for five example patients. Instructions to guide human authors through the process of developing similar synopses were written and revised based on feedback from clinicians. In parallel, a trained data scientist (S.B.) created prompts with the LLM (Lllama-3 8B) for data, information, knowledge and wisdom, and ran the prompts ten times each for the five example patients and five additional training patients. An investigator then rated the AI-produced synopses’ accuracy, synthesis, and conciseness (see Synopsis Rating section for more details) and provided guiding notes. The prompts were revised and re-run until at least eight of the ten resulting iterations were judged as successful on at least two of these three criteria for each AI-produced synopsis.

### Synopsis Rating

For the LLM to human comparison, we asked reviewers to grade each pair of synopses in a blinded, randomized fashion. Training consisted of presentation of the same example synopses with example scores. Human-and LLM-generated synopses were rated by clinicians on four of the fifteen PDQI constructs: accuracy, synthesis, conciseness, and usefulness, all rated with a 5-point Likert scale. 10% of the synopses were rated twice, with a total of 512 duplicated ratings (8 patients x 4 synopses x 2 creators x 4 ratings x 2 raters). Raters were presented with the source data (discharge notes and accompanying structured data) and with the data, information, knowledge, or wisdom synopsis to be rated.

### Analysis

We calculated descriptive statistics for patient demographics, clinical characteristics, and source data (note length and number of structured concepts). Human- and LLM-generated synopses were characterized according to length and, through the application of cTakes version 4.0,^38^ the number of relevant clinical concepts included.

We analysed the adequacy of the prompt engineering through graphic comparison of variability with a goal of reaching consistent performance.

Rater agreement was assessed with Krippendorff’s Alpha and the Intraclass Correlation. The former measures the likelihood precise agreement is due to chance, while the latter measures rater reliability.

Non-inferiority analyses were completed to compare the succinctness, accuracy, usefulness, and synthesis of the LLM-generated synopses compared to the human-generated synopses generated on the same patient. For analysis purposes, the ratings were dichotomized: a synopsis was successful according to a given construct if the assigned rating was a 4 (mostly) or 5 (very). **Clinically significant** differences were set at 5%, where 1 of 20 synopses didn’t meet criteria. **Statistical analysis** compared differences between LLM and human response through the Fisher’s exact test, with the McNemar test for uneven rates of false positives and false negatives. These analyses were performed for the data, information, knowledge, and wisdom synopses separately.

## Results

Sixty-four patients were summarized by both the LLM and Humans, with 8 double summarized (12%). Table 1 provides descriptive statistics for the patients. In all, synopses were written on 64 patients; 61% were male; 80% white; and the most common diagnoses were hypertension, ischemic heart disease, diabetes, and cancer. Compared to the MIMIC3 distribution overall, the selected sample was slightly younger (MIMIC3 median age 66); slightly more male (MIMIC3 56% male).

**Table 1.** Descriptive Statistics for included patients.

| Characteristic | N (%) |
| --- | --- |
| Total patients | 64 (100%) |
| Female Sex | 25 (39%) |
| Race/Ethnicity |  |
| Black/African American | 8 (13%) |
| Other | 2 (3.1%) |
| Unknown | 1 (4.7%) |
| White | 51 (79.6%) |
|  | Median (Q1, Q3) |
| Age | 58 (50, 68) |
| Chronic Conditions | N (%) |
| Ischemic Heart Disease | 19 (30%) |
| Diabetes | 19 (30%) |
| Autoimmune diseases | 1 (1.6%) |
| Chronic Kidney Disease | 8 (13%) |
| Heart failure | 6 (9.4%) |
| Dementia | 1 (1.6%) |
| Cancer, excluding skin cancer | 17 (27%) |
| Mood disorders, including depression, anxiety, bipolar | 8 (13%) |
| Stroke | 13 (20%) |
| Anemia | 22 (34%) |
| Hyperlipidemia | 15 (23%) |
| Hypertension | 37 (58%) |
| Hyperthyroidism | 6 (9.4%) |
| Osteoporosis | 2 (3.1%) |
| Osteoarthritis | 2 (3.1%) |

**Table 2.** Descriptive statistics for the synopses; numbers reported are median counts. All differences between Clinician- and LLM-generated summaries were significant under the Wilcoxon rank sum test with an alpha of 0.05 (p<0.001 in all cases).

|  | Clinician generated | LLM generated |
| --- | --- | --- |
| **Data** |  |  |
| Total words | 87 | 63 |
| Concepts included | 22 | 16 |
| Sentences | 4 | 3 |
| **Information** |  |  |
| Total words | 70 | 95 |
| Concepts included | 15 | 24 |
| Sentences | 4 | 5 |
| **Knowledge** |  |  |
| Total words | 74 | 86 |
| Concepts included | 15 | 21 |
| Sentences | 3 | 3 |
| **Wisdom** |  |  |
| Total words | 66 | 96 |
| Concepts included | 10 | 16 |
| Sentences | 3 | 3 |

### Prompt engineering iterations

The initial and final prompts used are in the Supplement. Figure 2 displays the median and range of scores rated as successful out of 10 LLM generations across the 10 example patients. For Data, the median of scores met the 80% threshold, but scores as low as 30% were seen. For information, 2 of 3 ratings met criteria, with wide variation in succinctness on initial runs. For Knowledge, 2 of 3 met criteria, with synthesis both poorly performing (50% median success rate) and variable (0-100% range). Wisdom met all three criteria but had highly variable accuracy and succinctness. All prompts were changed to reflect this initial review.

### LLM and Human blinded comparison

Raters scored all synopses in a blinded fashion; across all 4 statistics, the % of successful scores was 76.9% for AI and 82.6% for humans. As shown in Figure 3, AI performed *at similar success levels to* humans in Accuracy across Data, Information, and Wisdom, with an 8% decreased success in Knowledge. The AI generally rated *much worse* in Succinctness than humans, with 19-36% more AI synopses failing Information, Knowledge and Wisdom. Data Synopses from the AI were judged more succinct. For synthesis, AI was *much worse* than humans for data; but similar in Information and Knowledge synopses. Eight percent more AI-produced Wisdom synopses met success criteria than humans. For usefulness, the AI rated worse in Data, the same in Information and Knowledge, and much better in Wisdom (+11% success rate), with 75% of AI responses rated as useful compared to 76% for humans.

**Figure 2.**
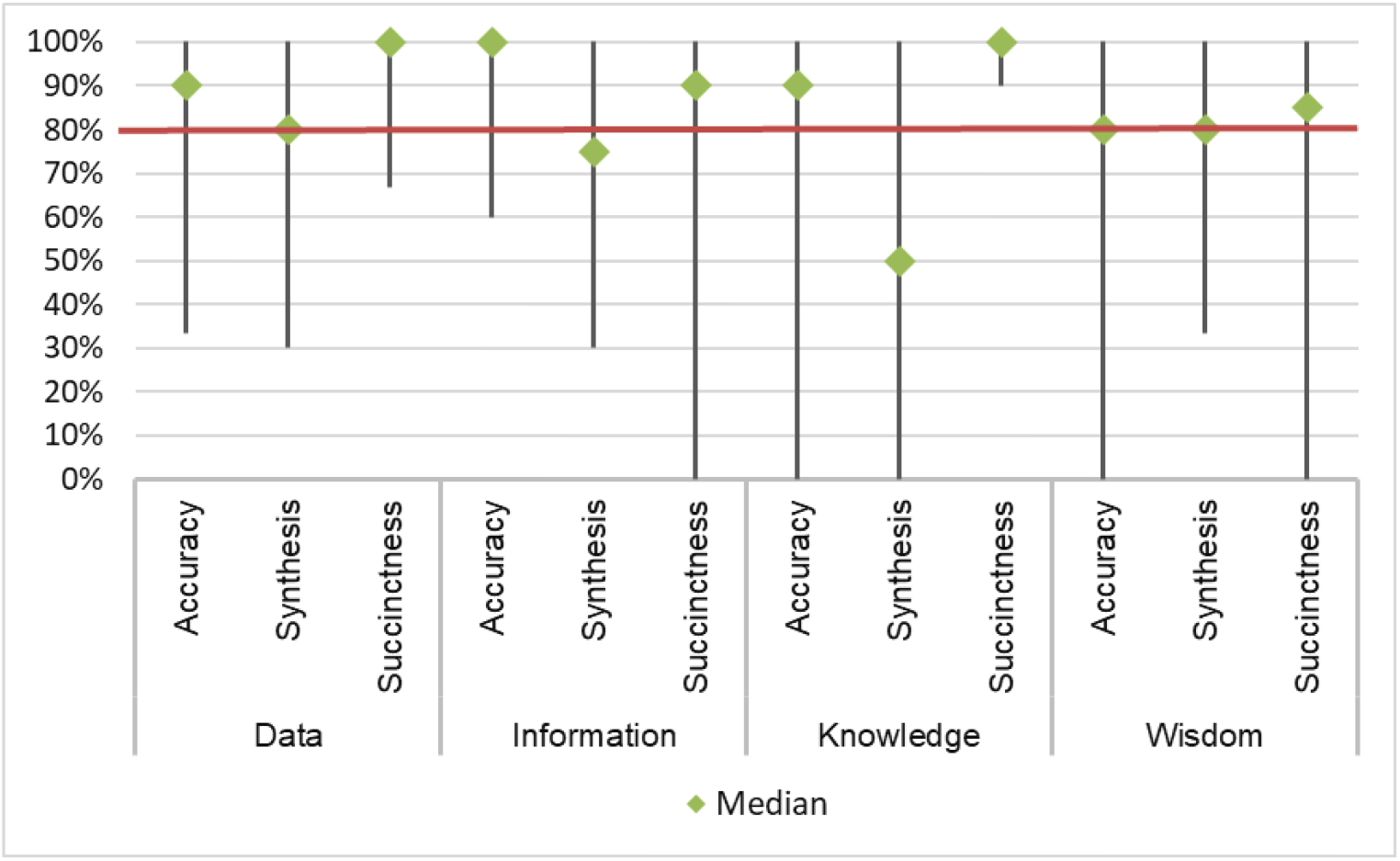
Median and variation in % rated 4 or 5 across categories for initial set (N=10)

**Figure 3.**
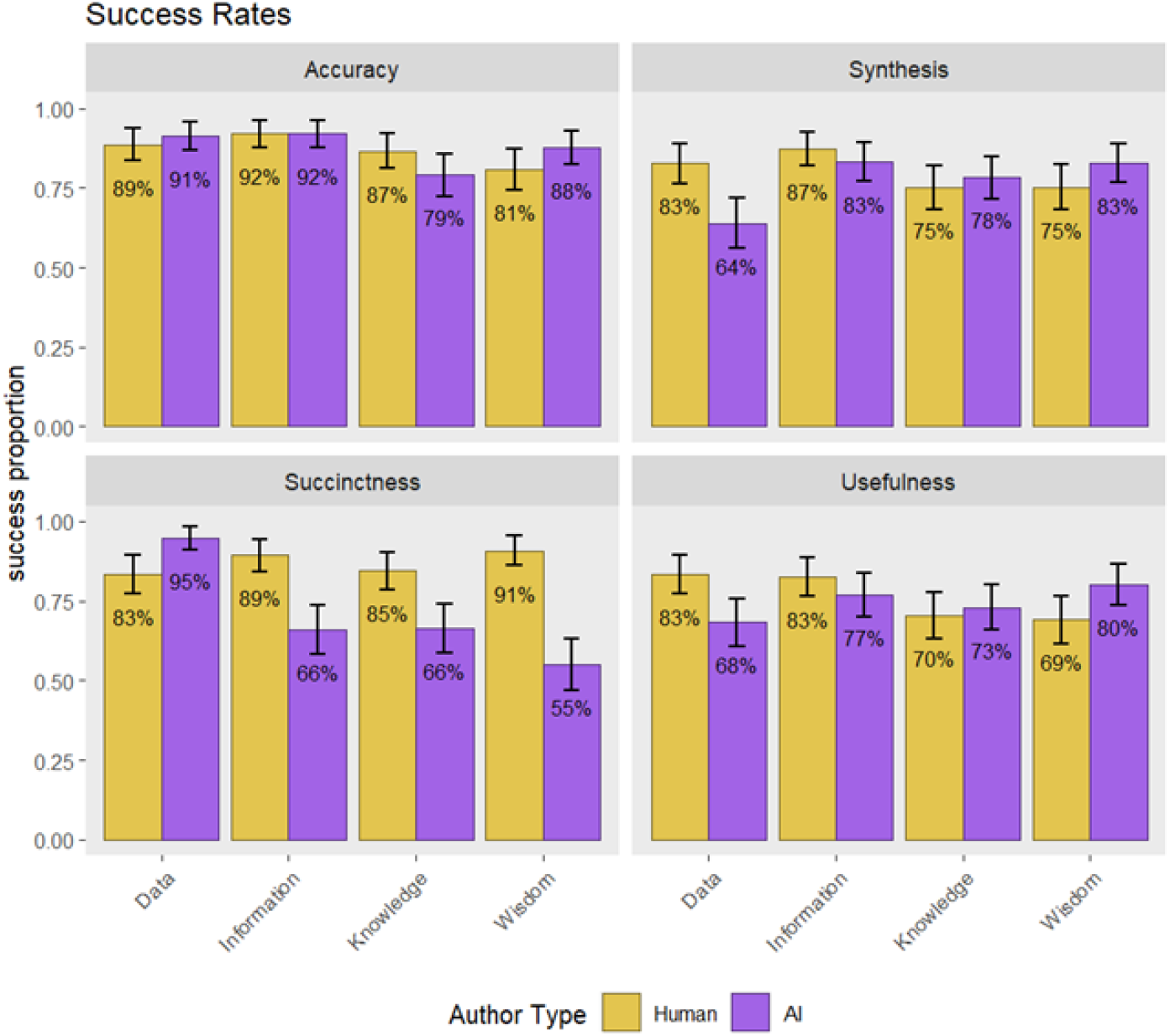
Success ratings (4 or 5) for AI and Human-produced synopses across Data, Information, Knowledge, and Wisdom Categories.

Agreement. Of the 10% of synopses that were double rated, Krippendorff’s Alpha was moderate (.224) for precise agreement – 3 raters had higher overall scores versus the other two.

However, the intraclass correlation for raters was high (.92), indicating overall reliability. Neither varied for the ratings for human and AI-generated synopses.

## Discussion

The advent of Generative AI capabilities, especially for text production, have enabled a new set of health documentation and communication capabilities, including text generation, explanation, and summarization.^23-26,28-32^ In this study, we successfully completed a rigorous protocol for benchmarking a clinically relevant data summarization tool. Our results showed that iterative fine tuning of a generative AI model produced brief summaries of patient needs after hospitalization that were non-inferior to human-created ones in synthesis and usefulness and potentially superior in more holistic, wisdom-based reflections.

The succinctness of its summarizations varied, with Data synopses by the AI rated highly succinct, but all other categories were not succinct. There seemed to be a trade-off between succinctness and other categories – synthesis and usefulness were both rated much lower for the AI than humans in data, while less succinct AI synopses rated more highly.

We were able to train this generalist Generative AI tool to generate results zero-shot fashion (albeit with chaining) and achieved high performance through careful prompt engineering. The creation and testing of prompts is a crucial aspect of fine-tuning any model; for ours, it required several iterations with repeated rating to achieve reasonable scores. These prompts are available in the Appendix.

The **value** of this work is three-fold. First, it demonstrates the relative performance of LLMs in these common, time-consuming summarization tasks as compared to humans across a set of standard measures. Second, it provides a robust, reproducible protocol for others to use as they train, evaluate, and implement LLMs; we provide the full protocol, including the prompts, instructions, and examples and invite others to replicate our experiment and benchmark their own LLMs. Third, and perhaps most usefully, it parses summarization into different levels based on a key framework originally from nursing – Data-Information-Knowledge-Wisdom – that will allow others to better specify their specific solution and parse errors in a more granular fashion. It also facilitates chaining – moving from less to more integrative tasks through serial steps – which are a particular strength of LLMs. Thus, it provides a useful model on which others may build.

Summarization may be one of the greatest features of LLMs: pulling out the most common ideas and themes from extensive documents (16, 19-20,22). The burden both of producing and reading documentation is one being addressed in several areas, including through a program intended to reduce documentation burden by 25% in five years.^39^ The utility of accurate and thoroughly synthesized synopses for patients with significant illness burden or after care transitions is particularly high.

Limitations. The data we used was not wholly representative of data for patients with complex needs; we limited available data and MIMIC-III is a subset of high-risk patients. Performance may vary across different populations. LLMs are known to hallucinate; although we scored the synopses on accuracy, automation bias may have led raters to accept confident statements as truth. This was most likely in the Knowledge section, where the LLM quoted key guidelines that it had previously consumed: raters may not have checked these. Adding quantitative checks for hallucinations and additional LLM verification layers are the next steps to address this weakness. The PDQI scores we used had moderate agreement on scores, indicating variability in preference for synopses; this variability was the same for humans and AI.

Future work. Expanding beyond discharge summaries is a crucial next step for outpatient visit synopses. Adams et al. (2021) reviewed the nature of hospitalization summaries by constructing a dataset of 109,000 patients; they find discharge summaries to be highly concise, somewhat comprehensive, and with unique organization compared to other notes.^40^ Thus, summarizing other note types into synopses may prove difficult. This is especially poignant given Kripalani’s findings that discharge summaries lack many elements that may require follow-up.^41^

## Conclusions

We created a protocol to benchmark LLMs for synopsis creation after hospital discharge using a Data-Information-Knowledge-Wisdom framework. We tested the protocol on an open-source LLM and blind-rated its outputs versus those of humans, achieving similar success rates for usefulness, worse scores for succinctness, and better scores in the Wisdom task.

## Data Availability

All data produced in the present study are available upon reasonable request to the authors. The data on which the work was done is available on Physionet.org

https://physionet.org/content/mimiciii/1.4/

## References

1. Chan C-L, You H-J, Huang H-T, Ting H-W. Using an integrated COC index and multilevel measurements to verify the care outcome of patients with multiple chronic conditions. BMC Health Services Research. 2012/11/19 2012;12(1):405. doi:10.1186/1472-6963-12-405

2. Clerc P, Boyer V, Haramburu F, Fourrier-Reglat A, Le Breton J. Identifying High-Risk Medication Prescriptions to Prevent Potentially Severe Adverse Drug Events in Primary-Care Patients with Chronic Multimorbidities: The Polychrome Study. Journal of Pharmacy and Pharmacology. 2020;8:35–43.

3. Lehnert T, König H-H. Auswirkungen von Multimorbidität auf die Inanspruchnahme medizinischer Versorgungsleistungen und die Versorgungskosten. Bundesgesundheitsblatt-Gesundheitsforschung-Gesundheitsschutz. 2012;55(5):685–692.

4. Maciejewski ML, Mi X, Curtis LH, Ng J, Haffer SC, Hammill BG. Few disparities in baseline laboratory testing after the diuretic or digoxin initiation by medicare fee-for-service beneficiaries. Circulation: Cardiovascular Quality and Outcomes. 2016;9(6):714–722.

5. Schoen C, Osborn R, How SK, Doty MM, Peugh J. In Chronic Condition: Experiences Of Patients With Complex Health Care Needs, In Eight Countries, 2008: Chronically ill US patients have the most negative access, coordination, and safety experiences. Health affairs. 2008;27(Suppl1):w1-w16.

6. Porter J, Boyd C, Skandari MR, Laiteerapong N. Revisiting the time needed to provide adult primary care. Journal of general internal medicine. 2023;38(1):147–155.

7. Rule A, Bedrick S, Chiang MF, Hribar MR. Length and Redundancy of Outpatient Progress Notes Across a Decade at an Academic Medical Center. JAMA Netw Open. Jul 1 2021;4(7):e2115334. doi:10.1001/jamanetworkopen.2021.15334

8. Downing NL, Bates DW, Longhurst CA. Physician burnout in the electronic health record era: are we ignoring the real cause? : American College of Physicians; 2018. p. 50–51.

9. Cutler DM, Ly DP. The (paper) work of medicine: understanding international medical costs. Journal of Economic Perspectives. 2011;25(2):3–25.

10. Kim P, Daly JM, Berry-Stoelzle MA, et al. Prognostic Indices for Advance Care Planning in Primary Care: A Scoping Review. J Am Board Fam Med. Mar-Apr 2020;33(2):322–338. doi:10.3122/jabfm.2020.02.190173

11. Edwards ST, Dorr DA, Landon BE. Can Personalized Care Planning Improve Primary Care? Jama. Jul 4 2017;318(1):25–26. doi:10.1001/jama.2017.6953

12. Tajirian T, Stergiopoulos V, Strudwick G, et al. The influence of electronic health record use on physician burnout: cross-sectional survey. Journal of medical Internet research. 2020;22(7):e19274.

13. McDuff D, Schaekermann M, Tu T, et al. Towards accurate differential diagnosis with large language models. arXiv preprint arXiv:231200164. 2023;

14. Graves JR, Corcoran S. The study of nursing informatics. Image: the journal of nursing scholarship. 1989;21(4):227–231.

15. Targowski A. From data to wisdom. Dialogue & Universalism. 2005;15

16. Matney S, Brewster PJ, Sward KA, Cloyes KG, Staggers N. Philosophical approaches to the nursing informatics data-information-knowledge-wisdom framework. Advances in Nursing Science. 2011;34(1):6–18.

17. Cato KD, McGrow K, Rossetti SC. Transforming clinical data into wisdom: Artificial intelligence implications for nurse leaders. Nurs Manage. Nov 2020;51(11):24–30. doi:10.1097/01.NUMA.0000719396.83518.d6

18. Hultman GM, Marquard JL, Lindemann E, Arsoniadis E, Pakhomov S, Melton GB. Challenges and Opportunities to Improve the Clinician Experience Reviewing Electronic Progress Notes. Appl Clin Inform. May 2019;10(3):446–453. doi:10.1055/s-0039-1692164

19. Payne TH, Keller C, Arora P, et al. Writing Practices Associated With Electronic Progress Notes and the Preferences of Those Who Read Them: Descriptive Study. J Med Internet Res. Oct 6 2021;23(10):e30165. doi:10.2196/30165

20. Nijor S, Rallis G, Lad N, Gokcen E. Patient Safety Issues From Information Overload in Electronic Medical Records. J Patient Saf. Sep 1 2022;18(6):e999–e1003. doi:10.1097/pts.0000000000001002

21. Alqenae FA, Steinke D, Keers RN. Prevalence and Nature of Medication Errors and Medication-Related Harm Following Discharge from Hospital to Community Settings: A Systematic Review. Drug Saf. Jun 2020;43(6):517–537. doi:10.1007/s40264-020-00918-3

22. Zaretsky J, Kim JM, Baskharoun S, et al. Generative artificial intelligence to transform inpatient discharge summaries to patient-friendly language and format. JAMA network open. 2024;7(3):e240357–e240357.

23. Van Veen D, Van Uden C, Blankemeier L, et al. Adapted large language models can outperform medical experts in clinical text summarization. Nature medicine. 2024;30(4):1134–1142.

24. Park Y-J, Pillai A, Deng J, et al. Assessing the research landscape and clinical utility of large language models: A scoping review. BMC Medical Informatics and Decision Making. 2024;24(1):72.

25. Andrew A. Potential applications and implications of large language models in primary care. Family Medicine and Community Health. 2024;12(Suppl 1)

26. Pivovarov R, Elhadad N. Automated methods for the summarization of electronic health records. Journal of the American Medical Informatics Association. 2015;22(5):938–947.

27. Pivovarov R, Coppleson YJ, Gorman SL, Vawdrey DK, Elhadad N. Can Patient Record Summarization Support Quality Metric Abstraction? AMIA Annu Symp Proc. 2016;2016:1020–1029.

28. Liang J, Tsou C-H, Poddar A. A novel system for extractive clinical note summarization using EHR data. 2019:46–54.

29. Devlin J. Bert: Pre-training of deep bidirectional transformers for language understanding. arXiv preprint arXiv:181004805. 2018;

30. Lee J, Yoon W, Kim S, et al. BioBERT: a pre-trained biomedical language representation model for biomedical text mining. Bioinformatics. 2020;36(4):1234–1240.

31. Šuvalov H, Laur S, Kolde R. Information Extraction from Medical Texts with BERT Using Human-in-the-Loop Labeling. Caring is Sharing–Exploiting the Value in Data for Health and Innovation. IOS Press; 2023:831–832.

32. Peng Y, Yan S, Lu Z. Transfer learning in biomedical natural language processing: an evaluation of BERT and ELMo on ten benchmarking datasets. arXiv preprint arXiv:190605474. 2019;

33. Meta-Llama-3-8B. Meta. https://huggingface.co/meta-llama/Meta-Llama-3-8B

34. Stetson PD, Bakken S, Wrenn JO, Siegler EL. Assessing electronic note quality using the physician documentation quality instrument (PDQI-9). Applied clinical informatics. 2012;3(02):164–174.

35. Goldberger AL, Amaral LA, Glass L, et al. PhysioBank, PhysioToolkit, and PhysioNet: components of a new research resource for complex physiologic signals. Circulation. Jun 13 2000;101(23):E215–20. doi:10.1161/01.cir.101.23.e215

36. Gorina Y, Kramarow EA. Identifying chronic conditions in Medicare claims data: evaluating the Chronic Condition Data Warehouse algorithm. Health Serv Res. Oct 2011;46(5):1610–27. doi:10.1111/j.1475-6773.2011.01277.x

37. Mullenbach J, Pruksachatkun Y, Adler S, et al. CLIP: a dataset for extracting action items for physicians from hospital discharge notes. arXiv preprint arXiv:210602524. 2021;

38. Savova GK, Masanz JJ, Ogren PV, et al. Mayo clinical Text Analysis and Knowledge Extraction System (cTAKES): architecture, component evaluation and applications. J Am Med Inform Assoc. Sep-Oct 2010;17(5):507–13. doi:10.1136/jamia.2009.001560

39. Levy DR, Withall JB, Mishuris RG, et al. Defining Documentation Burden (DocBurden) and Excessive DocBurden for All Health Professionals: A Scoping Review. Applied Clinical Informatics. 2024;15(05):898–913.

40. Adams G, Alsentzer E, Ketenci M, Zucker J, Elhadad N. What’s in a Summary? Laying the Groundwork for Advances in Hospital-Course Summarization. Proc Conf. Jun 2021;2021:4794–4811. doi:10.18653/v1/2021.naacl-main.382

41. Kripalani S, LeFevre F, Phillips CO, Williams MV, Basaviah P, Baker DW. Deficits in communication and information transfer between hospital-based and primary care physicians: implications for patient safety and continuity of care. Jama. 2007;297(8):831–841.

