## Supplemental File for "Can Electronic care planning using AI Summarization Yield equal Documentation Quality? (EASY eDocQ)"

**Appendix A.1 Data-Information-Knowledge-Wisdom Framework**

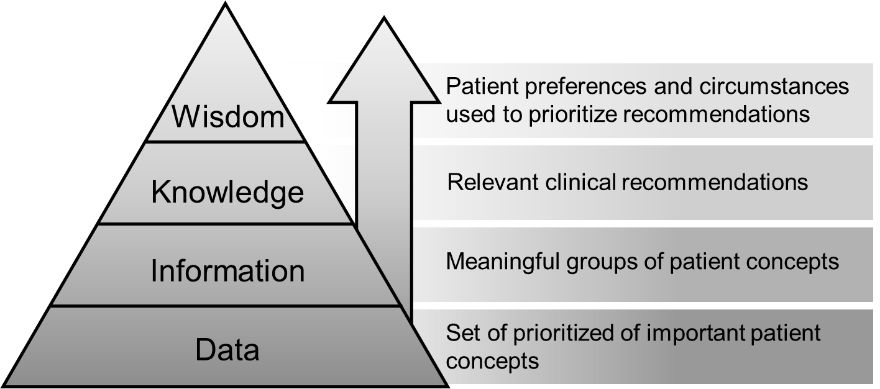
We used the Data-Information-Knowledge-Wisdom framework to define levels of summarization that are used in care planning preparation. Data is defined as the key data elements needed to consider for current care planning. These become information when placed in the patient’s context, usually through grouping by condition or need. Knowledge incorporates guidelines or other recommendations about appropriate, evidence-based treatment for these conditions. This step requires outside information not usually available directly from the chart. Finally, Wisdom attempts to take a holistic approach that incorporates the values and preferences of patients and their health trajectory, social situation, and other issues.

**Appendix A.2 Human Synopsis Generation Instructions**

We recruited six physician participants to generate synopses of de-identified discharge notes from PhysioNet’s MIMIC-III database, including one attending physician, one fellow, and four residents. First, each participant was instructed to complete a series of required tasks and trainings to gain access to the MIMIC-III database (see Aim 1.1: Human synopsis generation training materials). The study team then selected 80 discharge summaries from the MIMIC-III database, assigning each physician participant between 12 and 16 summaries to produce synopses. We selected summaries that included a patients with multiple chronic conditions, where conditions were selected from the Chronic Condition Warehouse (eTable 1). These were subdivided into requiring substantial follow-up or variable follow-up; patients needed to have at least one of the former.

**eTable 1. Chronic Conditions selected for knowledge assessment**

| **Requiring substantial follow-up** | **Requiring variable follow-up** |
| --- | --- |
| Ischemic Heart Disease | Anemia |
| Diabetes | Hyperlipidemia |
| Autoimmune diseases | Hypertension |
| Chronic Kidney Disease | Hypothyroidism |
| Heart failure | Osteoporosis |
| Dementia | Osteoarthritis |
| Cancer, excluding skin cancer |  |
| Parkinson’s |  |
| COPD/Asthma |  |
| Mood disorders |  |
| Stroke |  |

Each discharge summary was assigned a unique document ID (DocID) to track assignments and link synopses to their corresponding summaries. Participants received their assigned discharge summaries via individual, secure folders emailed directly to them. The study team also developed a Qualtrics survey to introduce the project, outline its objectives, and explain the Data, Information, Knowledge, and Wisdom (DIKW) framework in detail, along with step-by-step instructions for completing the synopsis-generation task. Each level of the DIKW framework included specific instructions that participants followed when crafting their synopses –

- **Data**: Create a summary for this patient by data domain – the most important diagnoses, the most important labs, vitals, medications, and other information. Don’t combine the information at this stage. Be very brief and focus on only the most important pieces of information; do not summarize the entire note. This is a summarization step that you may not write down, but may use to guide your note – just ensuring the problems, the medications, the labs/studies, and other data you feel are important are listed.
- **Information**: Create a 3-sentence summary for this patient, focused on putting together the data with the most important patient context, e.g., combining key information about this patient with the data to generate key areas that will likely to be addressed at the next outpatient visit for an internal medicine provider. This is a common step when you may be presenting information by condition or need or summarizing in an assessment. Try to avoid making the plan, just highlight the information and its relevance in an organized fashion.
- **Knowledge**: Based on your understanding of the core guidelines for these conditions, create a 3-sentence summary that contains recommendations from these guidelines and how they apply to this patient. This step is akin to a plan by condition with relevant information kept within it.
- **Wisdom**: Focusing on the expressed values and preferences of this patient, and any relevant behavioral and social context, create a 3-sentence summary that aligns the medical data and information about this patient with the relevant guideline recommendations with the values, preferences, behavioral and social context. This step is only sometimes completed, essentially during reflection of the patient’s overall trajectory and our broader consideration of their needs, goals, values and preferences. This sometimes may be akin to preparing for Advance Care Planning or similar conversations, but other times may be thoughtful prioritization and reflection given their course so far. This may be difficult and is okay to be brief if there isn’t enough context.

Each physician participant received a unique Qualtrics survey link allowing them to pause and return to the task at their convenience, with their progress automatically saved. Participants were instructed to write four synopses for each discharge summary, following the DIKW framework prompts. To familiarize participants with the process, they were provided an example discharge summary along with a sample synopsis before beginning. The Qualtrics survey then guided participants through the synopsis-writing task, providing the DIKW prompts and dedicated space on separate pages for each discharge summary to ensure clarity and organization. Participants entered their synopses directly into the Qualtrics survey and submitted their responses to the study team upon completing the assigned discharge summaries.

At the end of the survey, participants were asked a series of questions about which step they considered most important for clinical care, the perceived difficulty of the task, how they chose to order and group information within their synopses, and any surprises they encountered during the process.

Part A.2.1: Human synopsis generation training materials

Before initiating the synopsis generation task, all physician participants were instructed to complete a series of required trainings to gain access to PhysioNet resources, including the de-identified discharge summaries from the MIMIC-III database. These trainings ensured compliance with PhysioNet’s data use requirements and allowed participants to work with the de-identified discharge summaries.

The study team compiled all required training material instructions and emailed them to each physician participant. The training requirements included:

1. **Account Setup on PhysioNet**:
   a) Go to [PhysioNet’s homepage](https://physionet.org/) to create a user account
2. **Collaborative Institutional Training Initiative (CITI) Training Modules and Instructions:**
   a) Next, visit [PhysioNet’s CITI course instructions](https://physionet.org/about/citi-course/) to view instructions on how to complete the required courses.
3. **Add an Affiliation**:
   a) First, log in to CITI using your OHSU credentials.
   b) Next, you are required to Massachusetts Institute of Technology (MIT) as an affiliate institution within your OHSU CITI account. Follow PhysioNet’s instructions on how to add MIT before completing the required CITI modules.
4. **Complete the required CITI modules:**
   a) After affiliating with MIT, you should add and complete the “Data or Specimens Only Research” and “Conflict of Interest” CITI modules. Be sure to download and save your completion certificates for each module.
5. **Uploading CITI Certificates to PhysioNet**:
   a) Once the CITI trainings are complete, log in to your PhysioNet account and navigate to Account Settings.
   b) Select the Training tab in the left menu and upload your completed CITI training certificates.
6. **Credentialing in PhysioNet**:
   a) Under Account Settings, choose the Credentialing tab and provide the required information.
      1. For the Referral Section, list a supervisor or colleague who is an approved PhysioNet user as a reference.
         1. This individual will receive and email notification to approve you.
      2. In response to the field regarding research goals and datasets, include this in your description: *“We are conducting research on automated methods for note summarization and consolidation using large language models. Our study leverages fine-grained annotations in the CLIP dataset to evaluate algorithm performance, using both the CLIP and MIMIC-III datasets.”*
7. **Complete the Data Use Agreement (DUA)**:
   a) The final step is to sign a DUA for MIMIC-III database access. Follow this link to complete your DUA: [MIMIC-III DUA](https://physionet.org/sign-dua/mimiciii/1.4/).

Each physician participant was instructed to notify the study team upon completing the required trainings. After the study team confirmed completion, the synopsis generation task was administered according to the methods outlined in Aim 1: Human Synopsis Generation Overview.

Appendix A.3 LLM Synopsis Generation Prompts

The LLM was trained through iterative draft prompts. Although the intent was to create a zero shot version, we started by asking the LLM to create 10 different versions with the same input to understand consistency. Initial prompts started with embedded data (see example in Version 1 below), then were adapted to incorporate lists of conditions and the most recent discharge summary.

Version one of a prompt template:

--------------------------------------------------------------------------------------------------------

Task: I will provide some information about a patient being seen in an outpatient clinic by a <provider type> and you will generate brief summaries. Note that these build on each other.

First Note: "Patricia Noelle, a 76 year old female, is having an office visit . Patricia has diabetes, chronic kidney disease (Stage 3), Congestive Heart Failure, Dyslipidemia, and Anxiety. Her medications are Insulin NPH/Reg 70/30 at 45 units twice per day; Lisinopril 40mg; Simvastatin 40mg; Furosemide 20mg; and Aspirin 81mg; Here CBGs range from 140-250, mostly around 180s;

Cr over last year (~ roughly every 3 months) ranged from 1.4, 1.6, 1.5, 1.9 today; and A1c over last 2 years started at 7.8, 8.1, 8.4, and is 9.1 today.

Patricia is concerned about her weight, which is up 5 pounds, her increased shortness of breath, which comes on with minimal activity. Reviewing her diet and activity goals, she notes the pandemic has worsened her anxiety and caused her to eat more comfort food."

Q: Create a summary for this patient by data domain – the most important diagnoses, the most important labs, vitals, medications, and other information. Don’t combine the information at this stage. Be very brief and focus on only the most important pieces of information; do not summarize the entire note.

A:[INFO_ONE]

Q: Create a 3-sentence summary for this patient, focused on putting together the data with the most important patient context, e.g., combining key information about this patient with the data to generate key areas that will likely to be addressed at the next outpatient visit for an internal medicine provider.

A:[INFO_TWO]

Task: The following are recommendations from evidence-based guidelines.

Q:Leveraging these guidelines, create a three-sentence summary that contains recommendations from these guidelines and how they apply to this patient.

Task:

Q:Focusing on the expressed values and preferences of this patient, and any relevant behavioral and social context, create 3 sentence summary that aligns the medical data and information about this patient with the relevant guideline recommendations with the values, preferences, behavioral and social context.

Q:

Second Note: "Patricia has fallen and broken a hip and is transferred from the hospital for s/p hip fracture repair and rehab in SNF. Her glipizide has been held, she comes to SNF with glargine 40 and sliding scale insulin.

All other home medications restarted at discharge to SNF with the additional medications for pain:

•	Oxycodone 5 mg q4 hr PRN

•	Tylenol 1000 mg every 8 hrs scheduled

•	ASA 81 mg BID x 6 weeks for DVT prophylaxis. On day #2 at SNF, she had orthostatic vitals done:

BP lying: 144/76

Standing: 122/64

Fasting CBGs have been ~130

Pre-prandial CBGs have been 150-250 and she is getting between 2-6 units of SS insulin at mealtime. Discharge problems include

•	Type 2 diabetes complicated by retinopathy, peripheral vascular disease, and CKD (eGFR 27)

•	Hypertension

•	Congestive Heart Failure

•	Dyslipidemia

•	Anxiety

•	Osteoporosis

•	Orthostatic Hypotension

•	High fall risk

"

Q: Given this new note, what are the most important pieces of information to include in a two-sentence summary of this patient's current status? Do not summarize the entire note, only include the most important information.

A:[INFO_TWO]

-------------------------------------------------

A variant:

Task: Read a series of clinical notes, and answer questions about each one paying special attention to pieces information that are present in the current note but were not present in the previous notes.

First Note: "Patricia Noelle, a 76 year old female, is having an office visit . Patricia has diabetes, chronic kidney disease (Stage 3), Congestive Heart Failure, Dyslipidemia, and Anxiety. Her medications are Insulin NPH/Reg 70/30 at 45 units twice per day; Lisinopril 40mg; Simvastatin 40mg; Furosemide 20mg; and Aspirin 81mg; Here CBGs range from 140-250, mostly around 180s;

Cr over last year (~ roughly every 3 months) ranged from 1.4, 1.6, 1.5, 1.9 today; and A1c over last 2 years started at 7.8, 8.1, 8.4, and is 9.1 today.

Patricia is concerned about her weight, which is up 5 pounds, her increased shortness of breath, which comes on with minimal activity. Reviewing her diet and activity goals, she notes the pandemic has worsened her anxiety and caused her to eat more comfort food."

Q: Q: What are the most important pieces of information in the first note to include in a two-sentence summary of this patient's current status? Do not summarize the entire note, only include the most important information.

A:[INFO_ONE]

Second Note: "Patricia has fallen and broken a hip and is transferred from the hospital for s/p hip fracture repair and rehab in SNF. Her glipizide has been held, she comes to SNF with glargine 40 and sliding scale insulin.

All other home medications restarted at discharge to SNF with the additional medications for pain:

•	Oxycodone 5 mg q4 hr PRN

•	Tylenol 1000 mg every 8 hrs scheduled

•	ASA 81 mg BID x 6 weeks for DVT prophylaxis. On day #2 at SNF, she had orthostatic vitals done:

BP lying: 144/76

Standing: 122/64

Fasting CBGs have been ~130

Pre-prandial CBGs have been 150-250 and she is getting between 2-6 units of SS insulin at mealtime. Discharge problems include

•	Type 2 diabetes complicated by retinopathy, peripheral vascular disease, and CKD (eGFR 27)

•	Hypertension

•	Congestive Heart Failure

•	Dyslipidemia

•	Anxiety

•	Osteoporosis

•	Orthostatic Hypotension

•	High fall risk

"

Q: Q: What are the most important new pieces of information in the second note that the patient's primary care provider would need to know about this visit? Be very brief and focus on only the most important pieces of new information that were not in the first note; do not summarize the entire note.

A:[INFO_TWO]

Final version

Overall system prompt: “Generate focused summaries of a patient's note, following instructions about what topics should be focused on in each summary.”

**Data**: “Summarize this patient's most important diagnoses. Be very brief, do not include extraneous text, and produce output in the form of two or three short sentences, not lists. Focus solely on the specific data reported in the note- do not synthesize any information or draw any new conclusions. Include the most important diagnoses, relevant medical history, specific vital signs of note, or other relevant findings, including only those most essential to understand the patient's clinical picture. If the note mentions specific medications that are clinically relevant to the other conditions that you include in your summary, make sure to include those medications in your summary, as well as any other especially important medications that a doctor treating this patient would absolutely need to know about.”

**Information**: “Next, produce a higher-level summary of the note. This summary may combine specific data points from the note in order to produce a more informative summary; for example, by including information about which conditions prompted the use of a specific medication, by providing interpretation of specific very important or relevant vital signs, and by including a more general discussion of the patient's situation and medical context. If medications are included in the summary, be specific about which medication is being discussed; if it is not clear from the text of the note itself why a specific medication was prescribed, do not speculate as to the medication's purpose, but still include the medication in the summary. The summary should still be grounded entirely in facts present in the note itself, should not depend on any information not included in the note, and should still be very short and without extraneous text. The summary's audience will be medical practitioners, so acronyms and abbreviations should be used if they help make the summary more concise or informative.”

**Knowledge**: “Based on your understanding of the core guidelines for these conditions, create a three-sentence summary that contains recommendations from these guidelines and how they apply to this patient. Your reader is a clinician, so use as much jargon and as many abbreviations as necessary. Do not include any extraneous text or commentary.”

**Wisdom**: “Next, generate a very high-level 3-sentence summary of the patient's overall condition and prognosis going forward. The summary should the medical data and information about this patient with the relevant guideline recommendations with any relevant information about the patient's values, preferences, and behavioral context; if there is no relevant information provided in the original note about these factors, the summary may list relevant questions that the patient, their family, and their providers should be considering at this time. The audience for this summary is other medical providers, so use whatever relevant technical vocabulary, jargon, or abbreviations that may be necessary for clarity and concision.”

Appendix A.4 Synopsis Judgment Task

Five of the six physician participants from the initial synopsis-writing task, one attending physician, one fellow, and three residents, participated in a subsequent judgment task to evaluate the human-generated synopses against AI-generated synopses. Participants rated each synopsis based on four criteria: Accuracy, Synthesis, Succinctness, and Usefulness, using a scale from 1 to 5 (where 1 indicates "not at all" and 5 indicates "extremely").

- **Accuracy –** the note is true. It is free of incorrect information.
- **Synthesis –** the note reflects the author’s understanding of the patient’s status and ability to develop a plan of care.
- **Succinctness –** the note is brief, to the point, and without redundancy.
- **Usefulness** – the note is extremely relevant, providing valuable information and/or analysis.

To administer the task, the study team randomly assigned each participant between 13 and 14 sets of synopses to judge, with the attending physician completing a total of 38 judgments. Each set comprised eight synopses: four human-generated and four AI-generated, both adhering to the DIKW framework prompts. Because some discharge summaries were double-assigned during the synopsis creation, certain AI-generated synopses were naturally rated twice. To ensure consistency, the study team also double-assigned the human-generated synopses for the same discharge summaries for evaluation.

Each participant received a personalized judgment task folder containing a spreadsheet with the synopses to judge, complete with their identifying DocID, as well as the corresponding discharge summary documents and instructions on how to complete the task (see Aim 3.1: Judgment Training Materials). Participants were blinded to the origin (human or AI) of each synopsis and were not assigned to judge any synopses for discharge summaries they had previously written synopses for. Judgments were recorded in the spreadsheet, with each synopsis evaluated on levels of Accuracy, Usefulness, Succinctness, and Synthesis.

Aim 3.1: Judgment task training materials

Before beginning the judgment task, the study team provided each physician participant with a document containing an introduction to the task and explicit instructions for completion. This document included the following information:

1. **Introduction:**

Thank you for participating in this judgment exercise. You, your colleagues, and a Large Language model (LLM) have created 76 sets of patient synopses. We aimed to produce concise summaries of patient status and care utilizing de-identified discharge notes from recently hospitalized individuals. You will evaluate a sample of 13 – 14 synopsis sets, each of which was written either by an LLM or by a clinician. Each synopsis you review should adhere to one of the levels of the data-information-knowledge-wisdom (DIKW) framework: **data** is individual elements; **information** integrates patient context with data; **knowledge** encompasses diagnoses, prognosis, and effective treatments; and **wisdom** incorporates values, preferences, and aligning care plans with patient wishes. **Page 2** has more information about each category and includes the prompts provided to the LLMs and clinicians to guide the generation of synopses. We expect that this exercise will take approximately 2 hours.

2. **Instructions:**
   a) Review the relevant discharge summaries.
   b) Evaluate a randomly generated set of human and LLM-generated synopses (the LLM and human synopses may be listed first or second without a clear pattern)
      - Utilize the discharge summaries as a reference for judging the synopses.
   c) Rate the synopses on the following attributes –
      - **Accuracy –** the note is true. It is free of incorrect information.
      - **Synthesis –** the note reflects the author’s understanding of the patient’s status and ability to develop a plan of care.
      - **Succinctness –** the note is brief, to the point, and without redundancy.
      - **Usefulness** – the note is extremely relevant, providing valuable information and/or analysis.
   d) Assign each attribute a judgement score between 1 – 5, where 1 = not at all and 5 = extremely.

The synopses you will evaluate are in an Excel spreadsheet located in your assignment folder. Record your evaluations in that Excel spreadsheet. Feel free to pause, save your progress, and return to this exercise as needed.

Here is an example of where the human and LLM-generated synopses, along with your judgements, will go within your spreadsheet –

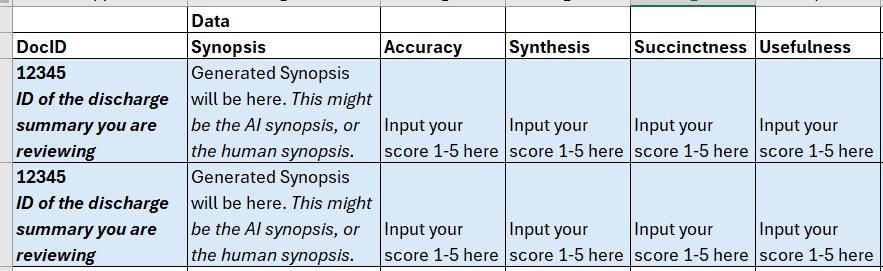

It's important to note these summaries gradually build upon each other. Don’t worry if the ratings appear to be challenging – just do your best to judge them based on your own experience and practice. If you absolutely cannot judge, write N/A. Recording N/A may be common in Wisdom-based examples where there is no obvious need described for person-centered care planning.

**Appendix A.4 DIKW Framework Instructions**

**These instructions were provided to the clinicians and the LLM who created the synopses. The purpose for including the instructions in this document is to provide reviewers with information about what each DIKW element is aiming to accomplish.**

**Data**

Objective*:* Create a summary for the data domain – the most important diagnoses, the most important labs, vitals, medications, and other information. Don’t combine information. Be very brief and focus on only the most important pieces of information; do not summarize the entire note. *This is a summarization step that you may not write down but may use to guide your note – just ensuring the problems, the medications, the labs/studies, and other data you feel are important are listed.*

**Information**

Objective: Create a 3-sentence summary for this patient, focused on putting together the data with the most important patient context, e.g., combining key information about this patient with the data to generate key areas that will likely to be addressed at the next outpatient visit for an internal medicine provider. *This is a common step when you may be presenting information by condition or need or summarizing in an assessment. Try to avoid making the plan, just highlight the information and its relevance in an organized fashion.*

**Knowledge**

Objective: Based on your understanding of the core guidelines for these conditions, create a 3-sentence summary that contains recommendations from these guidelines and how they apply to this patient. *This step is akin to a plan by condition with relevant information kept within it.*

**Wisdom**

Objective: Focusing on the expressed values and preferences of this patient, and any relevant behavioral and social context, create a 3-sentence summary that aligns the medical data and information about this patient with the relevant guideline recommendations with the values, preferences, behavioral and social context. *This step is only sometimes completed, essentially during reflection of the patient’s overall trajectory and our broader consideration of their needs, goals, values and preferences.* The best example of this kind of work is in holistic, person-centered care planning, which has several steps (see below). In the synopsis, you might see key references to these steps. Person-centered care planning is care that fits the person, often needing to adjust to a changing health trajectory where standard treatments may offer more burden and provide care discordant to evolving goals.
